# Follow-up Cervical Cancer Screening among Women living with HIV in Moshi Municipality, 2019-2024: Adherence and Predictors

**DOI:** 10.1101/2025.11.14.25340169

**Authors:** Dickson Doto, Laura Shirima, Blandina Mmbaga, Innocent H. Peter Ughh, Alex Mremi, Patricia Swai, Bariki Mchome

**Affiliations:** School of Public health, KCMC University, Tanzania; Obstetrics and Gynecology Department, Kilimanjaro Christian Medical Centre (KCMC), Tanzania; Pathology Department, Kilimanjaro Christian Medical Centre (KCMC), Tanzania; Kilimanjaro Clinical Research Institute (KCRI), Tanzania

**Keywords:** Cervical cancer screening, Follow-up, Adherence, Screening outcome, WLHIV

## Abstract

**Background:** Cervical cancer screening is a critical public health intervention that facilitate the early detection and treatment of precancerous cervical lesions before they progress to invasive cancer. The effectiveness of this intervention largely depends on timely and consistent adherence to recommended follow-up screenings. However, in Tanzania, the adherence to these crucial screenings and its associated predictors remain poorly understood.

**Objective:** To determine the proportion of WLHIV who adhered to a follow-up screening and its associated predictors among WLHIV who have undertaken their first screening between 2019 and 2022 in Moshi Municipality.

**Methodology:** This was a retrospective cohort study. Data were analyzed using STATA version 18.0. Descriptive statistics were employed to determine the proportion of adherence to follow-up screening. Multivariable logistic regression analysis was used to estimate adjusted odds ratios and 95% CI for the predictors of adherence.

**Results:** Among 3,076 WLHIV enrolled into the study, only 33.1% adhered to a follow-up screening. Among these, only 21.4% demonstrated good adherence. Also, adherence was higher in private healthcare facilities [AOR=2.55; 95% CI: 2.02–3.22], lower for WLHIV who had their first screening at an outreach facility [AOR=0.19; 95% CI: 0.12–0.29], higher for those who were diagnosed with abnormal screening outcome in their first screening [AOR=8.19; 95% CI: 4.11–16.34]. It also increases with age, and parity.

**Conclusion:** Adherence to follow-up screening among WLHIV in Moshi Municipality was low. Strengthening recall systems especially in public and outreach settings, along with targeted interventions for younger, low parity women and those with normal screening outcome in the first screening, is essential to improve adherence to these vital screenings

## Introduction

Cervical cancer screening is the crucial strategy in the fight against cervical cancer (1). It allows for the early detection and treatment of precancerous cervical lesions before they progress into invasive cervical cancer (2). Numerous studies have demonstrated that early cervical cancer screening combined with timely follow-up screenings can significantly reduce the burden of the disease (3,4). For instance, follow-up screening every 10 years can reduce the burden of cervical cancer by 40-60%, every 5 years by 85%, every 3 years by 91% and annually by 94% (5).

Much of the global disease burden is borne in sub-Saharan Africa where approximately 85% of cervical cancer cases and 90% of related deaths occurs (6). The disease is also much more prevalent in women living with HIV (WLHIV) (7,8). In response to this increased risk, WLHIV in Tanzania are eligible for their first cervical cancer screening as early as 25 years of age, compared to 30 years for women without HIV (9). Also, WLHIV are recommended to undertake a follow-up screening after a year compared to 3 years recommended for women with no HIV. Moreover, Visual Inspection with Acetic Acid (VIA) is currently the recommended cervical cancer screening method for women living with HIV (WLHIV) in the country. However, HPV DNA testing is under pilot implementation and is expected to be incorporated soon as an optional screening method (10).

In 2024, it was reported that 36.7% of WLHIV in Tanzania have undertaken the first cervical cancer screening (11). However, the proportion of those who adhered to a follow-up screening remains poorly understood. Also, several predictors including age, parity, marital status and screening outcomes in the first screening have been reported to be the predictors of adherence to a follow-up screening. However, these findings have been inconsistent across the globe and meanwhile no study conducted in the country or similar settings have conclusively determine those predictors (12–14). This study therefore aimed to determine the proportion of WLHIV who have adhered to a follow-up screening, the timing of their adherence and its associated predictors among WLHIV in Moshi Municipality.

## Materials and Methods

This was a retrospective cohort study, aiming to determine the proportion of adherence to a follow-up screening, the timing of their adherence, and its associated predictors among WLHIV who had undertaken their first screening between 2019 and 2022 in Moshi Municipality. For cervical cancer screening data, the study utilized data recorded in the National Health Management Information System (HMIS), which is locally known as *Mfumo wa Taarifa za Uendeshaji wa Huduma za Afya* (MTUHA). This system comprises both electronic and hardcopy registries, the electronic component contains aggregated summaries, while the hardcopy registries contain detailed, individual-level screening records. The system is designed to ensures consistency and standardization in data collection and reporting, thereby providing a reliable source of information for monitoring and evaluating the cervical cancer screening programme in the country (19). Meanwhile for HIV related data, the study utilized CTC2 database, and their information in that database were linked with their information in the cervical cancer screening registries, using their unique IDs.

Four facilities were purposively selected out of six which offer cervical cancer screening services in the Municipality. This includes Mawenzi Regional Referral Hospital, St. Joseph Hospital, Majengo Health Centre and *Chama cha Uzazi na Malezi Bora* (UMATI). The study area was purposively selected because the Kilimanjaro region, which includes Moshi Municipality, has consistently been ranked as the region with the highest prevalence of cervical cancer in the country (20). Additionally, the region is among the leading regions in the country in terms of the number of women screened for cervical cancer. So the selection of this study area ensured the inclusion of the most affected region by the disease in the country and guaranteed the availability of sufficient and reliable data to effectively address the study objectives (21).

Descriptive statistics were used to summarize the socio-demographic and clinical characteristics of the study participants. The results for categorical variables were presented using frequencies and proportions, while results for continuous variables were summarized using means and standard deviations. Moreover, STATA 18.0 software was used for data cleaning and analysis. The difference in the proportion of WLHIV who have adhered to a follow-up screening by independent variables was assessed using the Chi-square test or Fisher’s exact test (when expected cell counts were less than 5). Moreover, Multivariable logistic regression with robust standard errors was used to determine the odds ratios (OR) and their corresponding 95% confidence intervals (CI) for the predictors of adherence to a follow-up cervical cancer screening. The level of statistical significance was set at a p-value of <0.05.

Moreover, prior to model fitting, key assumptions of logistic regression were thoroughly assessed. To assess for the presence of outliers in the continuous variables - age and parity, boxplot diagrams were used. When evidence for outliers was detected, the winsorization technique was applied to remove the outliers from those variables, and as a result, new variable without outliers were then generated, categorized into distinct groups based on the previous studies, and subsequently used in the model fitting (22). Also, to assess for the presence of multicollinearity among the independent variables, the spearman correlation technique was employed. In cases where multicollinearity was evident, one of the highly correlated variables was excluded. Moreover, alternative to logistic regression models were not considered because the prevalence for cervical cancer screening in the Municipality was above 10%, which is the cut-off point for opting for such models.

Moreover, crude odds ratio (COR) was first estimated for each independent variable. Variables found to be statistically significant at p-value <0.05, those with potential confounding effect and known clinical/practical association with the dependent variable were included in the multivariable logistic regression model. Additionally, potential interaction effects among the independent variables were also assessed during the multivariable analysis. However, evidence for such effect was not found. Also, both forward and backward stepwise logistic regression techniques were used in the multivariable analysis to identify the most parsimonious model and estimate adjusted odds ratio (AOR) for the predictors. Moreover, model selection was guided by the Akaike Information Criteria (AIC), whereby the model with the lowest AIC value was selected as the final model.

## Results

A total of 3,076 participants were included in the study. Their mean age at the time of their first screening was 39.3 years, with the highest proportion, 1,121(36.4%), aged between 35 and 44 years. Most participants, 3,058(99.4%), were residing in Kilimanjaro region. Most participants, 2,557(83.1%), received their first screening at the healthcare facility rather than at an outreach facility. Additionally, the mean parity among the study participants was 2.7, with majority proportion, 2,213(71.9%), had between 2 and 4 parities at the time of the first screening.

Also, majority, 2,771(90.1%), had no other reproductive comorbidities. Most, 973(31.6%), were classified as having WHO HIV stage I at the time of the first screening. Moreover, regarding HIV viral load, majority, 2,360(76.7%), had undetectable levels (*<*50 copies/mm^3^) at the time of the first screening. Also, majority, 2,201(71.6%), had not yet developed Advanced HIV Disease (AHD) at the time of the first screening. Additionally, all of the screening procedures were performed by VIA method. Notably, all healthcare providers involved in the first screening, treatment of precancerous cervical lesions and follow-up screening were females **(Table 1)**.

**Table 1.**
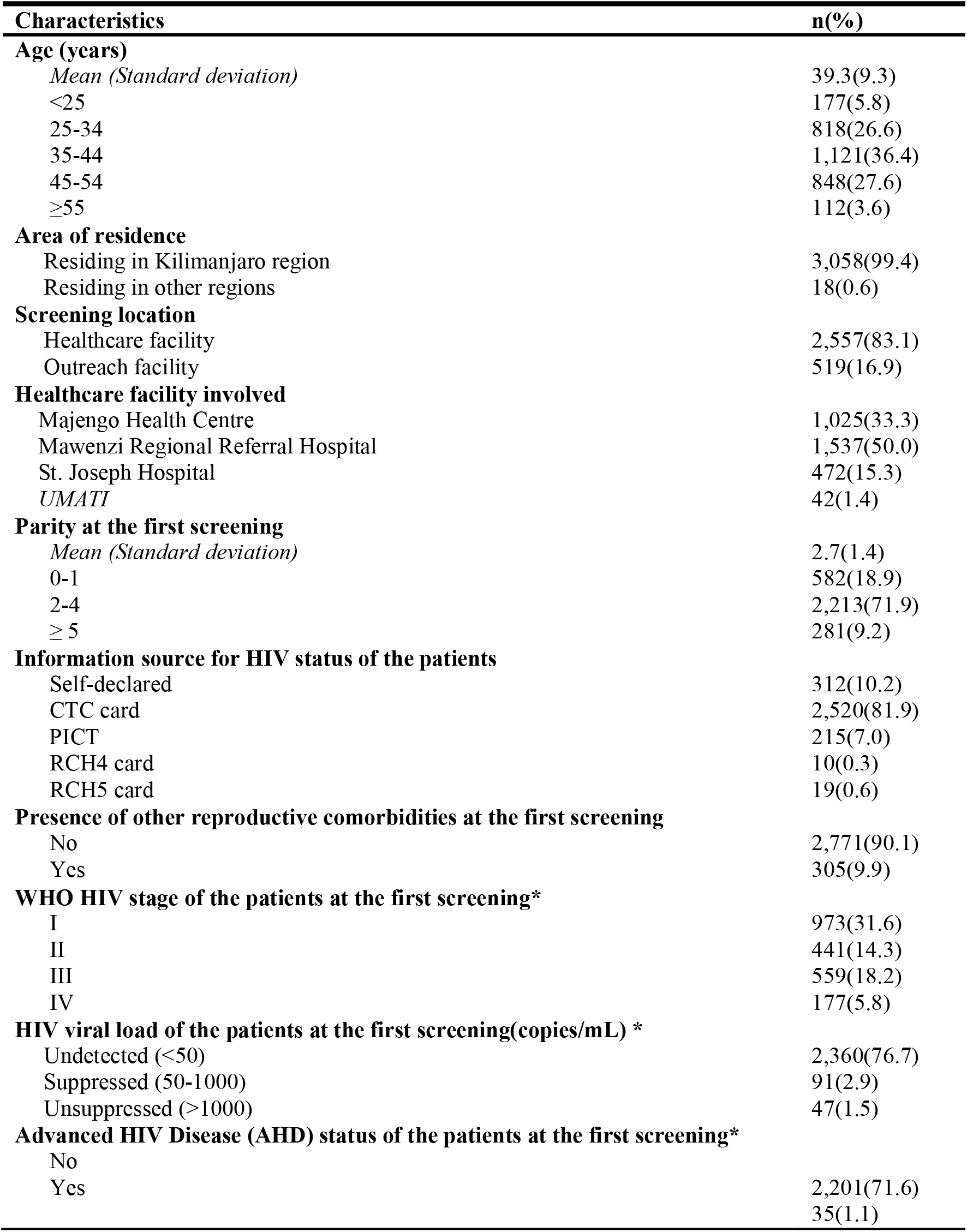
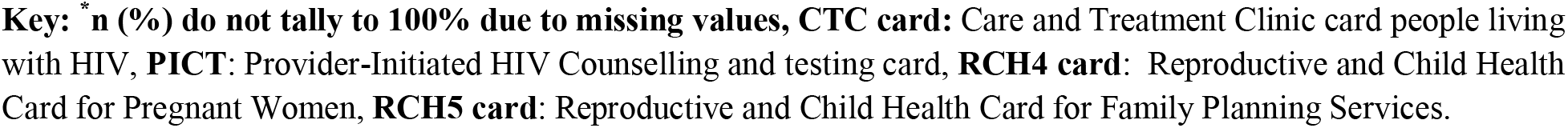
Socio-demographic and clinical characteristics of the study participants (n=3,076)

### Bivariate and multivariate analysis

Among 3,076 WLHIV who were enrolled into the study, 1,019(33.1%) [95%CI: 31.5% – 34.8%] adhered to a follow-up screening. Moreover, among those who adhered to a follow-up screening (n=1,019), only 218(21.4%) [95%CI: 18.9% – 24.0%] demonstrated good adherence. Others, 291(28.6%) [95%CI: 25.8% – 31.4%] demonstrated early adherence, 192(18.8%) [95%CI: 16.5% – 21.4%] demonstrated poor adherence whereas 318(31.2%) [95%CI: 28.4% – 34.2%] demonstrated extremely poor adherence to a follow-up screening.

A number of socio-demographic and clinical factors were statistically associated with adherence to a follow-up screening. These include, screening location, referral status of the healthcare facility, age, parity, presence of other reproductive comorbidities, screening outcome in the first screening, treatment modality of the precancerous lesions and AHD development status among the patients **(Table 2)**

**Table 2.**
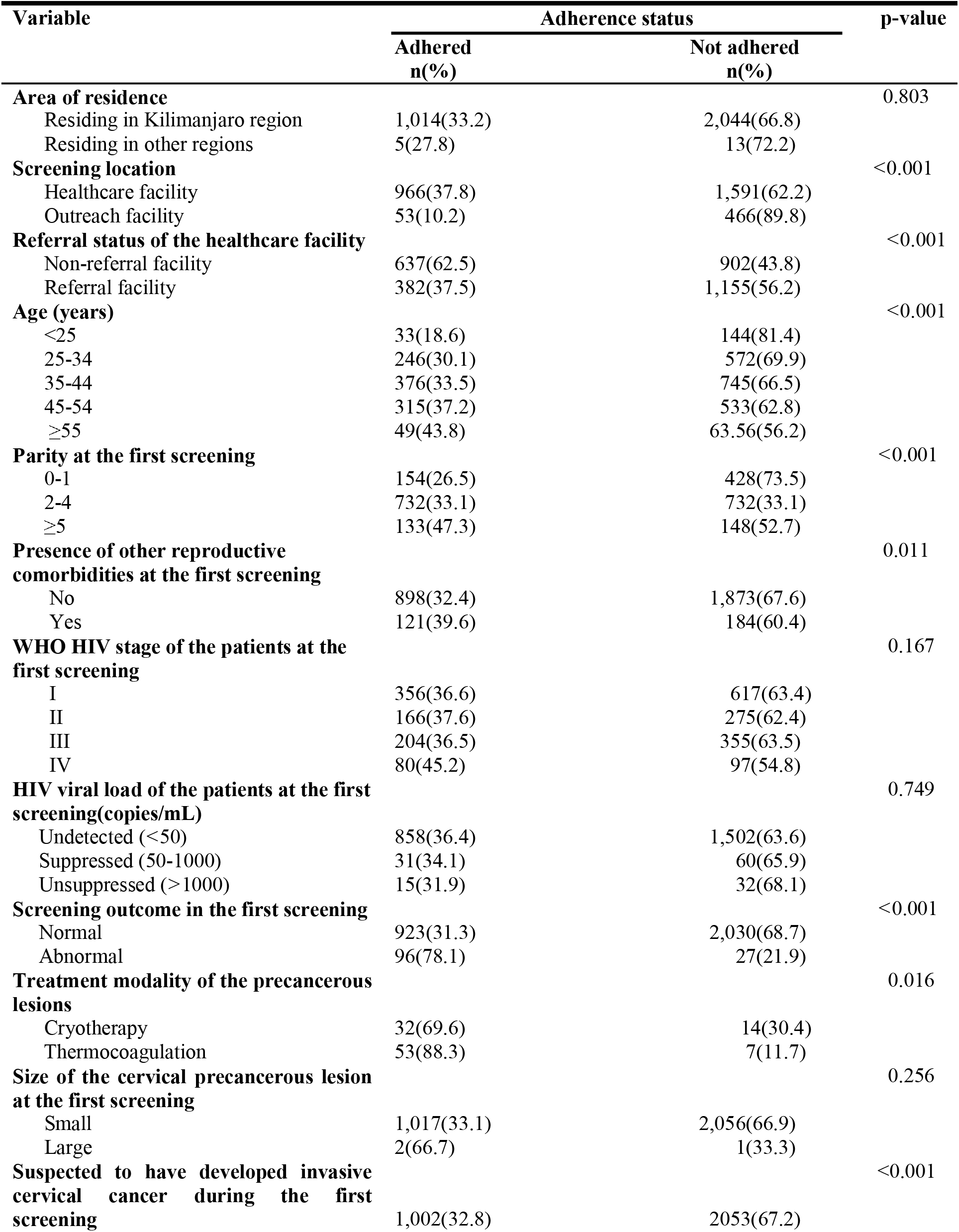
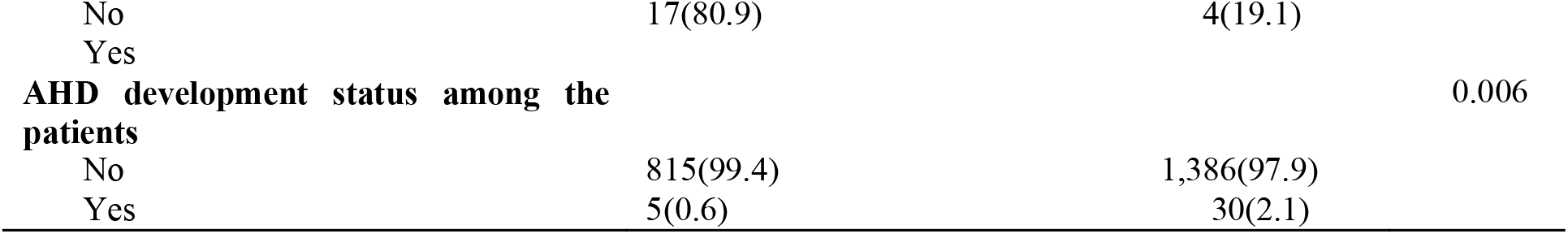
Adherence to a follow-up cervical cancer screening by socio-demographic and clinical factors among the study participants(n=3,076)

| Variable | Adherence status |  | p-value |
| --- | --- | --- | --- |
|  | Adhered<br>n(%) | Not adhered<br>n(%) |  |
| **Area of residence** |  |  | 0.803 |
| Residing in Kilimanjaro region | 1,014(33.2) | 2,044(66.8) |  |
| Residing in other regions | 5(27.8) | 13(72.2) |  |
| **Screening location** |  |  | <0.001 |
| Healthcare facility | 966(37.8) | 1,591(62.2) |  |
| Outreach facility | 53(10.2) | 466(89.8) |  |
| **Referral status of the healthcare facility** |  |  | <0.001 |
| Non-referral facility | 637(62.5) | 902(43.8) |  |
| Referral facility | 382(37.5) | 1,155(56.2) |  |
| **Age (years)** |  |  | <0.001 |
| <25 | 33(18.6) | 144(81.4) |  |
| 25-34 | 246(30.1) | 572(69.9) |  |
| 35-44 | 376(33.5) | 745(66.5) |  |
| 45-54 | 315(37.2) | 533(62.8) |  |
| ≥55 | 49(43.8) | 63.56(56.2) |  |
| **Parity at the first screening** |  |  | <0.001 |
| 0-1 | 154(26.5) | 428(73.5) |  |
| 2-4 | 732(33.1) | 732(33.1) |  |
| ≥5 | 133(47.3) | 148(52.7) |  |
| **Presence of other reproductive comorbidities at the first screening** |  |  | 0.011 |
| No | 898(32.4) | 1,873(67.6) |  |
| Yes | 121(39.6) | 184(60.4) |  |
| **WHO HIV stage of the patients at the first screening** |  |  | 0.167 |
| I | 356(36.6) | 617(63.4) |  |
| II | 166(37.6) | 275(62.4) |  |
| III | 204(36.5) | 355(63.5) |  |
| IV | 80(45.2) | 97(54.8) |  |
| **HIV viral load of the patients at the first screening(copies/mL)** |  |  | 0.749 |
| Undetected (<50) | 858(36.4) | 1,502(63.6) |  |
| Suppressed (50-1000) | 31(34.1) | 60(65.9) |  |
| Unsuppressed (>1000) | 15(31.9) | 32(68.1) |  |
| **Screening outcome in the first screening** |  |  | <0.001 |
| Normal | 923(31.3) | 2,030(68.7) |  |
| Abnormal | 96(78.1) | 27(21.9) |  |
| **Treatment modality of the precancerous lesions** |  |  | 0.016 |
| Cryotherapy | 32(69.6) | 14(30.4) |  |
| Thermocoagulation | 53(88.3) | 7(11.7) |  |
| **Size of the cervical precancerous lesion at the first screening** |  |  | 0.256 |
| Small | 1,017(33.1) | 2,056(66.9) |  |
| Large | 2(66.7) | 1(33.3) |  |
| **Suspected to have developed invasive cervical cancer during the first screening** |  |  | <0.001 |
|  | 1,002(32.8) | 2053(67.2) |  |
| No | 17(80.9) | 4(19.1) |  |
| Yes |  |  |  |
| **AHD development status among the patients** |  |  | 0.006 |
| No | 815(99.4) | 1,386(97.9) |  |
| Yes | 5(0.6) | 30(2.1) |  |

In multivariate analysis, after adjusting for the effect of other variables, a statistically significant association was found between adherence to a follow-up screening among the study participants and several independent variables. Specifically, participants who had their first screening at the private healthcare facility had 2.55 times higher odds of adhering to a follow-up screening compared to those who had their first screening at a public healthcare facility [AOR:2.55, 95%CI:2.02 – 3.22]. Also, participants who had their first screening at an outreach facility had 0.19 times lower odds of adhering to a follow-up screening compared to those who had their first screening at the healthcare facility [AOR:0.19, 95%CI: 0.12 – 0.29].

Moreover, adherence to a follow-up screening was increasing with age. For instance, participants who were aged 55 years and above, had 2.42 times higher odds of adhering to a follow-up screening compared to those who were aged below 25 years [AOR:2.42, 95%CI:1.21 – 4.84]. Also, adherence to a follow-up screening was increasing with parity. For instance, participants who had 5 children and above, had 1.67 times higher odds of adhering to a follow-up screening compared to those who had none or one child [AOR:1.67, 95%CI:1.11 – 2.49]. Also, participants who had abnormal screening outcome in the first screening, had 8.19 times higher odds of adhering to a follow-up screening compared to those who had normal screening outcome in the first screening [AOR:8.19, 95%CI:4.11 – 16.34]. Also, participants who had developed AHD, had 0.28 times lower odds of adhering to a follow-up screening compared to those who had not developed AHD [AOR: 0.28, 95%CI: 0.09 – 0.66] **(Table 3)**.

**Table 3.**
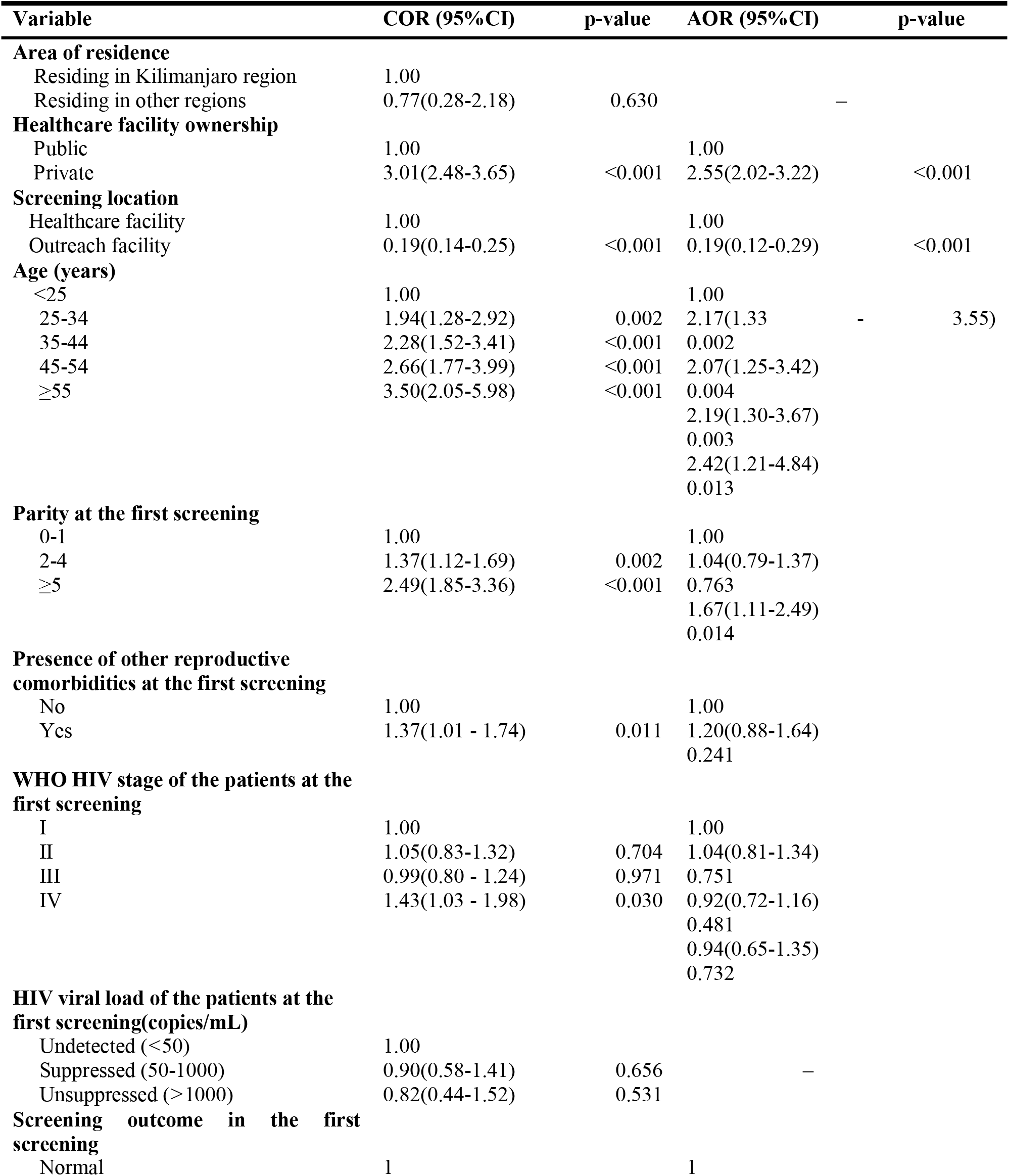
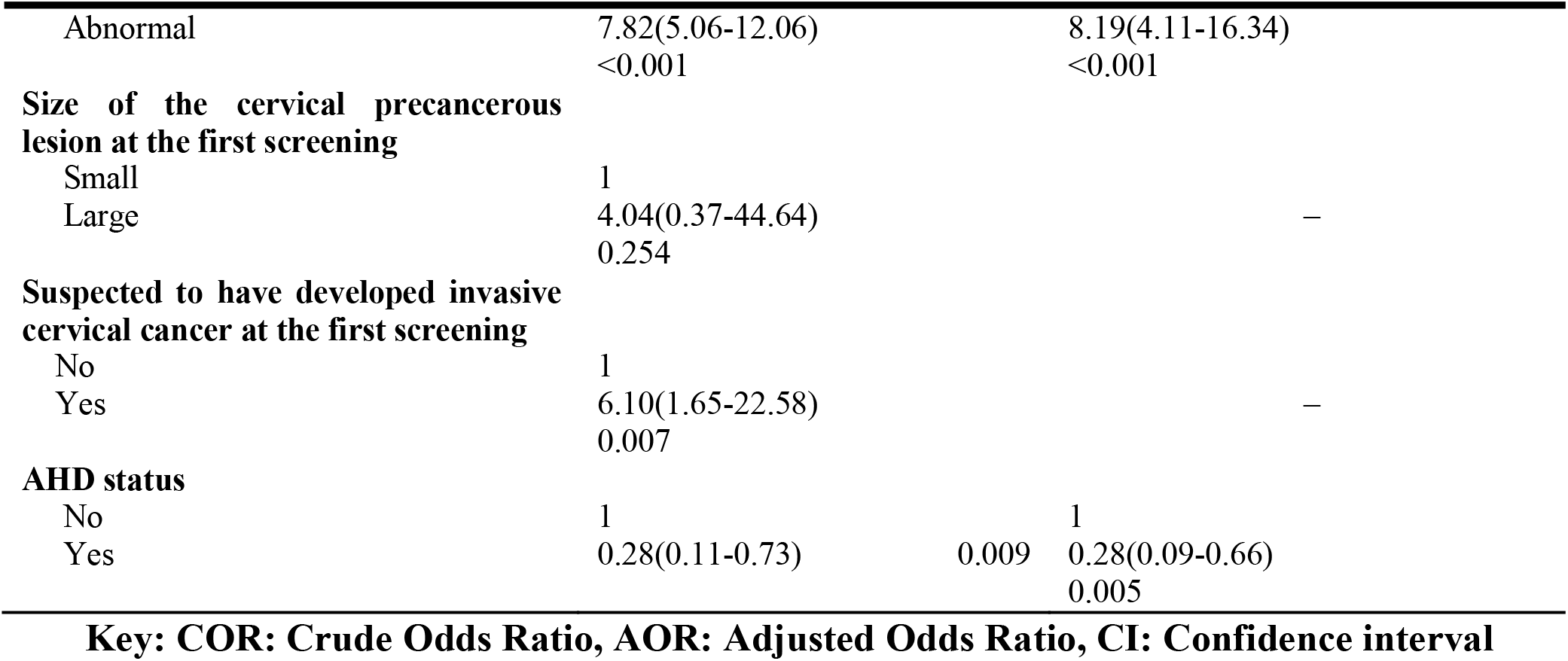
Multivariable analysis for predictors to a follow-up cervical cancer screening among WLHIV who had undertaken the first screening between 2019 and 2022 in Moshi Municipality (n=3,076)

## Discussion

This study found that only 33.1% of WLHIV who had undertaken the first cervical cancer screening had adhered to a follow-up screening. This low adherence may be attributed to insufficient recall and follow-up efforts made by the healthcare providers within their respective facilities, as well as inadequate knowledge about cervical cancer among the participants, which may have limited their understanding of the importance of timely adherence to a follow-up screening (23,24). Nevertheless, these findings are consistent with a similar study conducted in Nigeria, which reported an adherence of 37.8% to a follow-up cervical cancer screening (25). Such consistency suggests that the challenges of adherence to a follow-up cervical cancer screening among WLHIV may be common in low-resource settings.

Moreover, the study also revealed that among WLHIV who had adhered to a follow-up screening, only 21.4% demonstrated good adherence. Others, 28.6% demonstrated early adherence, 18.8% demonstrated poor adherence, and 31.2% demonstrated extremely poor adherence. These findings indicate that the majority of the small proportion of WLHIV who adhered to a follow-up screening did not show up within the recommended time frame (20). This underscores the critical gaps in the implementation of the intervention strategies intended to promote cervical cancer screening program in the country. In contrast, a study conducted in Denmark reported different patterns, among women who had undertaken the first cervical cancer screening, 26% demonstrated good adherence, 46% demonstrated poor adherence, 24% had more follow-up than recommended and only 4% were completely non-adherent (26). These disparities highlight the need for context-specific interventions to address the barriers to timely adherence to a follow-up screening in low-resource settings.

The study also highlighted a strong association between the ownership of healthcare facilities and adherence to a follow-up screening among WLHIV. This finding underscores the existing disparities between private and public healthcare facilities in promoting and facilitating follow-up care. Specifically, WLHIV who received cervical cancer screening services from private healthcare facilities were more likely to adhere to a follow-up screening compared those who went to public healthcare facilities, possibly due to better resource availability, more effective patient tracking systems, shorter waiting times, and improved quality of care. In contrast, public health facilities may face challenges such as understaffing, limited recall system, and inadequate patient follow-up mechanisms, which contribute to lower adherence in the follow-up screening (27). These disparities highlight the need for targeted strategies aiming to strengthen follow-up screening within public health settings including improved patient communication system.

The study also revealed a strong association between the location where the first cervical cancer screening was undertaken and adherence to a follow-up screening among WLHIV. Specifically, WLHIV who undertake the first cervical cancer screening at the healthcare facility were significantly more likely to adhere to a follow-up screening compared to those who were initially screened at an outreach facility. This finding suggests that healthcare facility-based screenings may offer more structured follow-up mechanisms, including proper documentation, better follow-up appointment scheduling, and more effective communication with patients. In contrast, outreach settings, while essential for expanding accessibility to screening services, may face challenges in maintaining patient records, providing timely reminders, and ensuring continuity of care (28). These results highlight the importance of integrating strong follow-up systems into outreach programs or linking them effectively with fixed healthcare facilities to improve adherence to follow-up screening among WLHIV (29).

Moreover, the study also revealed an association between the age of the participants and adherence to follow-up screening among WLHIV. Specifically, the findings have showed that older WLHIV were more likely to adhere to a follow-up screening compared to their younger counterparts (30). This trend may reflect increased health awareness, risk perception, health-seeking behavior and frequent interaction with healthcare services among older WLHIV (31). Conversely, younger WLHIV may be experiencing barriers such as limited health knowledge, competing priorities, or lower perceived susceptibility to cervical cancer. These findings suggest the need for age-targeted health education and counseling strategies, particularly focusing on younger WLHIV, aiming to improve their adherence to a follow-up cervical cancer screening. Nevertheless, these findings are consistent with previous studies conducted in Ethiopia and Ivory Coast, which similarly reported higher adherence to follow-up cervical cancer screening among older WLHIV (32,33).

The study also found a significant association between parity and adherence to a follow-up cervical cancer screening among WLHIV. Specifically, the results showed that WLHIV with high parity were more likely to adhere to a follow-up screening than those with fewer or no parity (34). This relation may be explained by the fact that WLHIV with multiple births tend to have more frequent contact with maternal and child health services, which may enhance their exposure to health education, screening opportunities, and the overall importance of preventive healthcare. Furthermore, high parity may reflect greater life experience and a heighted awareness of reproductive health risks, including cervical cancer, and thus encouraging better health-seeking behavior (35). These findings highlight the importance of integrating cervical cancer education and screening promotion into maternal health services, especially for women with lower or no parity who may not routinely engage with healthcare systems. The results of this study are consistent with the findings from previous studies conducted in Ethiopia, Uganda and Zimbabwe (32,34,36).

The study also revealed a strong association between the screening outcome in the first cervical cancer screening and subsequent adherence to a follow-up screening among WLHIV. Specifically, WLHIV who were diagnosed with abnormal screening outcome in the first screening were more likely to adhere to a follow-up screening compared to those who their first screening outcome were normal. This trend may be attributed to increased concern of perceived vulnerability among WLHIV with abnormal findings, which may prompt them to take recommended follow-up actions more seriously (31). Additionally, the emotional impact of an abnormal screening outcome may also enhance motivation to engage with healthcare services, follow-up treatment protocols, and attend follow-up visits. In contrast, WLHIV with normal screening outcome might perceive themselves as being at low risk and therefore may not prioritize returning for a follow-up screening (37). These findings emphasize the need for tailored counseling and education immediately following both normal and abnormal screening outcomes, to ensure all women understand the importance of routine follow-up regardless of the first screening outcomes. These results are consistent with the findings from a study conducted in Zimbabwe, which also reported higher follow-up adherence to a follow-up screening among WLHIV who had abnormal screening outcome in their first screening (8).

The study also found a significant association between AHD development status and adherence to a follow-up screening among WLHIV. Specifically, WLHIV who had not developed AHD were more likely to adhere to a follow-up screening compared to those who had developed AHD at the time of the first screening. This pattern may be due to the fact that WLHIV without AHD are generally healthier and more able to attend scheduled appointments, while those with AHD may face additional health burdens, reduced morbidity, or competing clinical priorities that hinder their ability to adhere to a follow-up cervical cancer screening (38). This finding highlights the need for targeted support strategies to improve follow-up adherence among WLHIV with AHD.

### Strength of the study

The study involved participants over a relatively long-time span, that is, from 2019 to 2024, which allowed for a broader and more reliable assessment of trends and patterns in adherence to follow-up cervical cancer screening among WLHIV. This extended period enhances the relevance and generalizability of the findings. Additionally, the study had also a sufficiently long follow-up time for the participants, making it possible to evaluate adherence behavior and clinical outcomes with greater accuracy.

Another strength lies in the inclusion of the healthcare facilities with diverse profiles. The study covered various facility types, including health centres, a regional referral hospital, and both public and private institutions, allowing for meaningful comparisons across different healthcare delivery contexts. This diversity strengthens the representativeness of the study population and makes the findings more applicable across different settings.

Also, adherence to a follow-up screening among WLHIV was categorized into detailed and distinct categories, that is early, good, poor and extremely poor categories, rather than using a simple binary classification. This approach provided a more nuanced understanding of adherence behavior and revealed specific gaps in the timing of follow-up cervical cancer screening.

Moreover, the study incorporated a wide range of potential predictors of adherence to follow-up screening, including both socio-demographic and clinical variables in the study. This comprehensive inclusion allowed for a more robust analysis and better understanding of the key determinants influencing a follow-up cervical cancer screening among WLHIV.

### Limitations of the study

A key limitation of this study was its inability to track participants who were classified as non-adherent at the healthcare facility where they received their first screening, to determine whether they may have undergone a follow-up screening at another selected healthcare facility. Since the study relied solely on records from individuals’ healthcare facilities, it is possible that some participants may have sought follow-up screening services at another healthcare facility rather than where they undertake the first screening. This limitation may have led to misclassification of adherence status and, consequently, an underestimation of the true adherence estimates.

Another limitation of the study lies in the fact that the registry from which data were obtained did not capture certain important demographic variables, including educational attainment, marital status, employment status and ethnicity of the participants. This information could have provided a deeper understanding of the social and demographic factors influencing adherence to follow-up screening and screening outcomes, thereby enriching the overall analysis and interpretation of the findings.

## Conclusion

Adherence to a follow-up cervical cancer screening among WLHIV was found to be remarkably low, with only a third of WLHIV adhering to it. This highlights a critical gap in the continuity of cervical cancer screening program among WLHIV in the country. Moreover, predictors significantly associated with adherence to a follow-up cervical cancer screening among WLHIV included healthcare facility ownership, screening location, age, parity, AHD status and screening outcome in the first screening. These findings emphasize the need for targeted interventions focusing particularly on younger women, those with low parity, and women screened in public or outreach settings to improve adherence to a follow-up screening. This should also involve strengthening of recall systems, patient tracking mechanisms, and individualized counseling, particularly for women who had normal screening outcomes in their first screening. This will play a key role in improving the overall effectiveness of the national cervical cancer screening programme among WLHIV, who are the most affected by the disease.

### Recommendations

The Ministry of Health should provide clear directives to the healthcare facilities on strengthening recall and follow-up strategies to improve WLHIV’s adherence to the follow-up cervical cancer screening appointments. This can be achieved by using evidence from previous studies in other settings, such as integrating community health workers home visitations to remind WLHIV who have undergone the first screening to adhere to a follow-up screening. Special attention should be given to public healthcare facilities, where adherence to a follow-up screening was found to be particularly low.

The Ministry of Health should consider adopting multiple testing methods, such as combining VIA, Pap smear, and HPV DNA testing as standard practice for cervical cancer screening among WLHIV. This approach would help to reduce false-negative results which will lead to improved diagnostic accuracy and ensure early detection of precancerous cervical lesions.

Additionally, stakeholders in the health sectors including clinicians at facility levels should plan to implement age- and parity-targeted interventions aiming to specifically promote follow-up cervical cancer screening among younger WLHIV and those with low parity, who were found to have low adherence to a follow-up cervical cancer screening compared to their older women and those with high parity counterparts, respectively. These interventions may include focused health education, peer support groups, and individualized counseling to improve risk perception and health-seeking behavior.

Clinicians at facility levels should also strengthen their counseling efforts for WLHIV who receive normal screening outcome during their first cervical cancer screening. In doing so, healthcare providers should be trained and supported to deliver structured and emotionally engaging health education emphasizing that a normal screening outcome does not eliminate future risk. On the other hand, National guidelines for cervical cancer screening should incorporate a standardized feedback mechanism on their cervical cancer screening outcome which will ensures all WLHIV understand the importance of routine follow-up screening, regardless of their screening outcome in the first screening.

Moreover, the Ministry of Health in the country, should consider formally recognizing and establishing distinct categories of adherence to a follow-up cervical cancer among women in the country. This categorization will enhance the monitoring and evaluation of cervical cancer screening program by enabling more precise tracking of patient’s behavior over time, identifying gaps in a follow-up care, and facilitating the design of targeted interventions to improve timely adherence and overall program effectiveness.

## Supporting information

supplemental attachments

## Data Availability

All data produced in the present study are available upon reasonable request to the authors

## Ethical approval

Ethical approval for this study was obtained from the Ethical Review Committee of KCMC University, under clearance number PG 174/2024. In addition, permission to access data from the selected healthcare facilities in Moshi Municipality was granted by the respective authorities.

## Acknowledgement

We thank the administration of the Moshi Municipality and the selected healthcare facilities for their invaluable and cooperation during the data acquisition process.

## Funding

We give our sincere gratitude to the Kilimanjaro Clinical Research Institute (KCRI) for their financial support towards this work.

## Conflict of interest

The authors declare no conflicts of interest regarding the publication of this paper

## Notes

### Competing Interest Statement

The authors have declared no competing interest.

### Funding Statement

This study was funded by Kilimanjaro Clinical Research Institute (KCRI) under the NORA project

### Author Declarations

KCMC university research ethical committee gave ethical approval for this work

