## supplemental attachments for "Follow-up Cervical Cancer Screening among Women living with HIV in Moshi Municipality, 2019-2024: Adherence and Predictors": Data extraction sheet.docx

### **Appendix 5: Data extraction sheet**

Date	………………

Participant ID …………….

**SECTION A: Demographic characteristics (check where applicable)**

1. Age ……….

2. Area of residence .................

3. Marital status

a. Single

b. Married

c. Cohabiting

d. Widowed

e. Divorced

4. Education level

a. No formal education

b. Primary

c. Secondary

d. Tertiary

5. Occupation

a. Peasant

b. Housewife

c. Employed

d. Self-employed

e. Petty cash traders/small business

f. Businesswoman

g. Other ……….

6. Health insurance status

a. No

b. Yes

7. Parity	…....……….

**SECTION B: Clinical characteristics**

8. Screening location

a. At the healthcare facility
b. At an outreach facility

9. Referral status of the healthcare facility

a. Non-referral facility
b. Referral facility

10. Information source for HIV status of the patients

a. Self-declared
b. Care and Treatment Clinic card people living with HIV (CTC card)
c. Provider-Initiated HIV Counselling and testing card (PICT)
d. Reproductive and Child Health Card for Pregnant Women (RCH4 card
e. Reproductive and Child Health Card for Family Planning Services (RCH5 card)

11. Date of the first cervical screening …………….

12. Patient’s CTC2 number …………..

13. First cervical cancer screening outcome

a. Normal

b. Abnormal

14. Screening modality in the first cervical cancer screening

a. VIA

b. Pap smear

c. HPV DNA test

d. Other

15. Size of the cervical precancerous lesion at the first screening

a. Small

b. Large

16. Suspected to have already developed invasive cervical cancer during the first screening

a. No

b. Yes

17. AHD development status among the patients during the first screening

a. No

b. Yes

18. Screening modality in the follow-up cervical cancer screening

a. VIA

b. Pap smear

c. HPV DNA test

d. Other

19. Presence of other reproductive comorbidities at the first screening

a. No

b. Yes

20. Treatment modality of the cervical precancerous lesions after abnormal screening outcome in the first screening

a. Cryotherapy

b. LEEP

c. Other

21. Presence of comorbidities during the first screening

a. No

b. Yes

22. Type of comorbidity during the first screening

a. Hypertension
b. Diabetes
c. Other

23. The qualification of the healthcare provider in the first screening

a. A nurse
b. Assistant medical doctor
c. Medical doctor
d. Medical specialist i.e. obstetrician and/or gynecologist

24. The qualification of the healthcare provider in the treatment of cervical precancerous lesions (when applied).

a. A nurse
b. Assistant medical doctor
c. Medical doctor
d. Medical specialist i.e. obstetrician and/or gynecologist

25. Sex of the healthcare provider during the first screening

a. Female

b. Male

26. Sex of the healthcare provider during the first screening

a. Female

b. Male

27. Date of a follow-up screening ……………
