## supplemental attachments for "Follow-up Cervical Cancer Screening among Women living with HIV in Moshi Municipality, 2019-2024: Adherence and Predictors": Extra Appendices.docx

### **Appendix 6: Extra analyses**

**Figure 4: Adherence to a follow-up cervical cancer screening among the study participants (n=3,076)**

Adherence to a follow-up cervical cancer screening among the study participants (%)

|  | % |
| --- | --- |
| Not adhered | 33.1 |
| Not adhered | 66.9 |

**Figure 5: Levels of adherence to a follow-up cervical cancer screening among the participants (n = 3,076)**

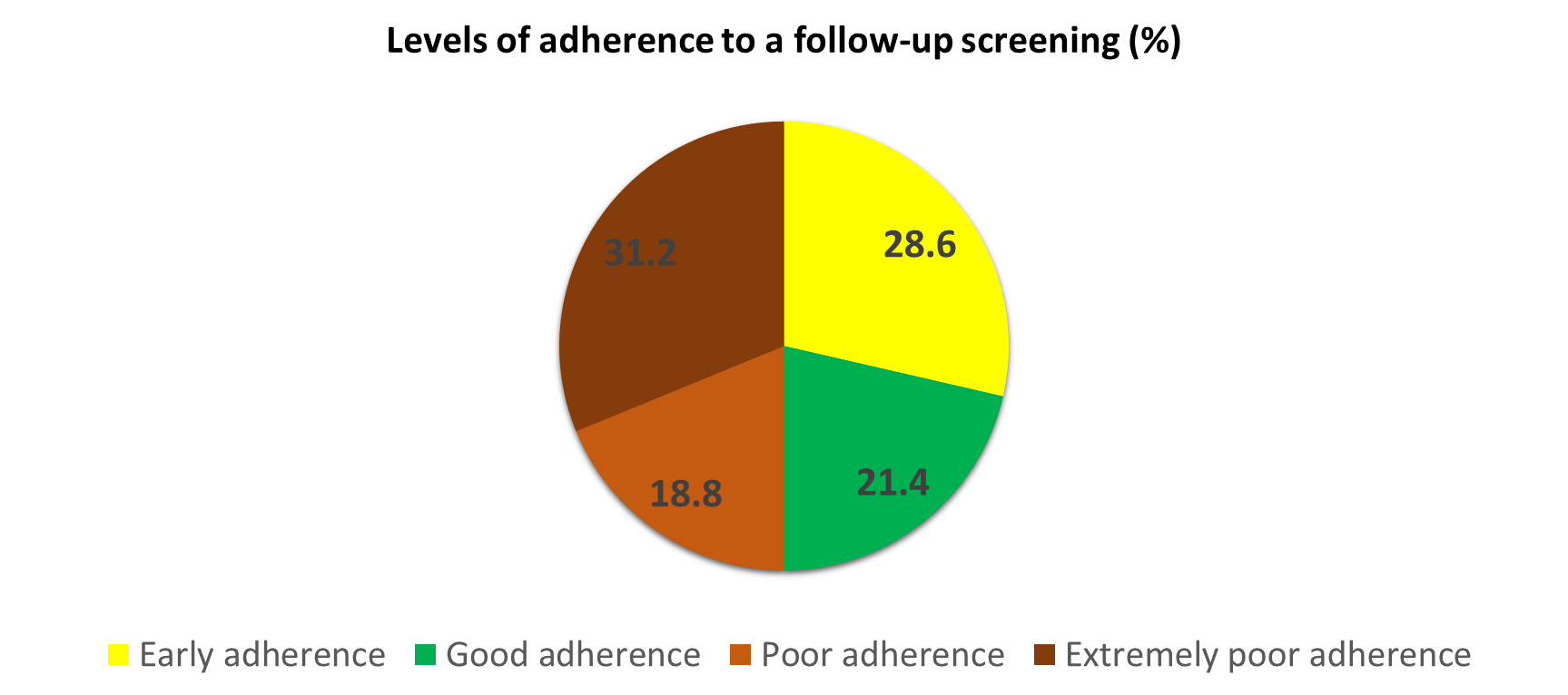

**Figure 6: Cervical cancer screening outcome among the study participants during their first screening (n=3,076)**

Cervical cancer screening outcome among the study participants during their first screening (%)

|  | % |
| --- | --- |
| Normal | 96 |
| Abnormal | 4 |

**Figure 7: Categories of abnormal screening outcomes during the first screening (n=3,076)**

Categories of abnormal screening outcomes during the first screening (%)

|  | % |
| --- | --- |
| Small lesions | 87.8 |
| Large lesions | 2.4 |
| Invasive cancer | 9.8 |

**Figure 8: Cervical cancer screening outcomes in the follow-up screening (n=1,019)**

Cervical cancer screening outcomes in the follow-up screening (%)

|  | % |
| --- | --- |
| Normal | 95.8 |
| Abnormal | 4.2 |
