## supplemental attachments for "Follow-up Cervical Cancer Screening among Women living with HIV in Moshi Municipality, 2019-2024: Adherence and Predictors": List of figures.docx

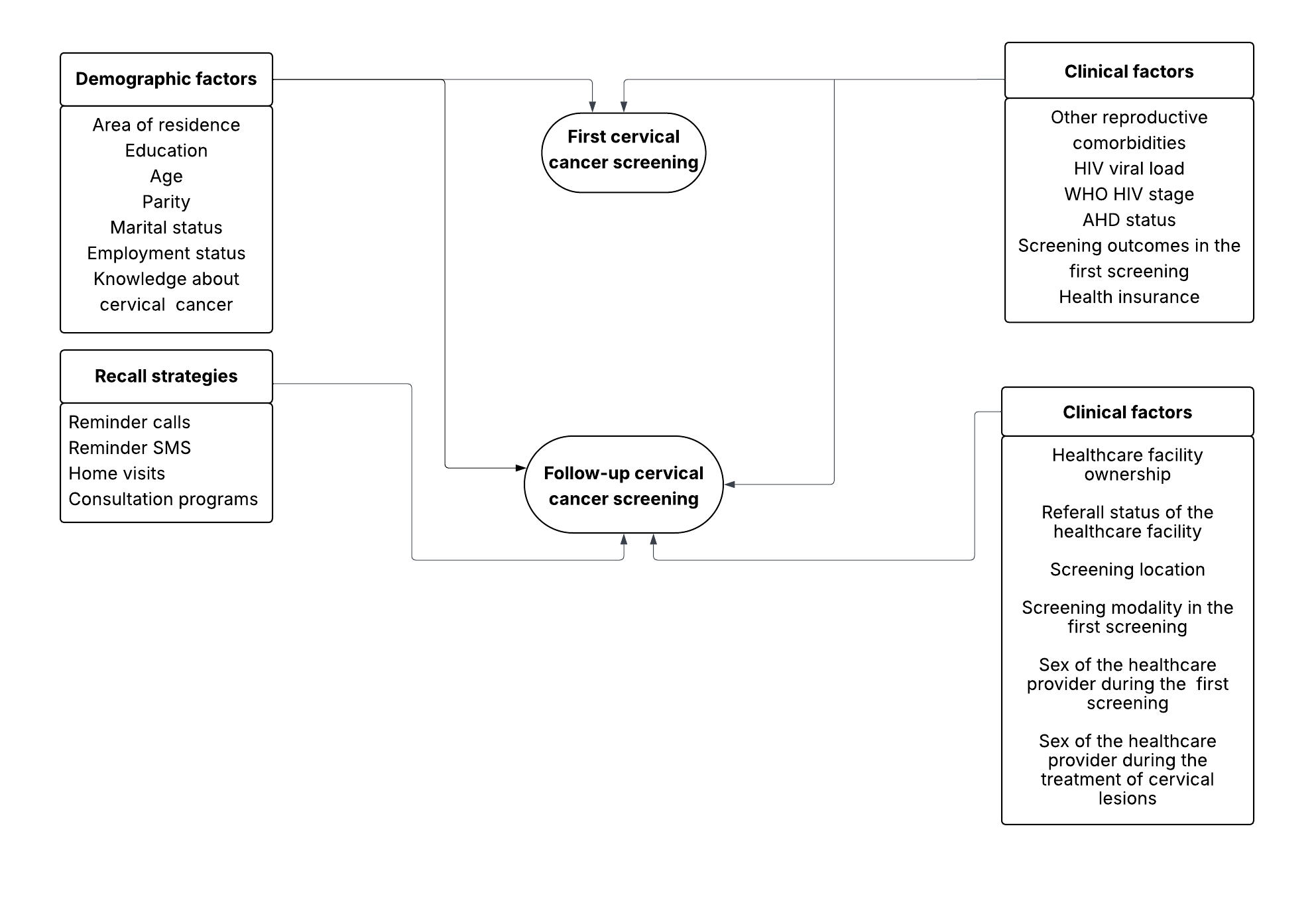
**Figure 1: Conceptual framework for a follow-up cervical cancer screening among WLHIV** (15–17)

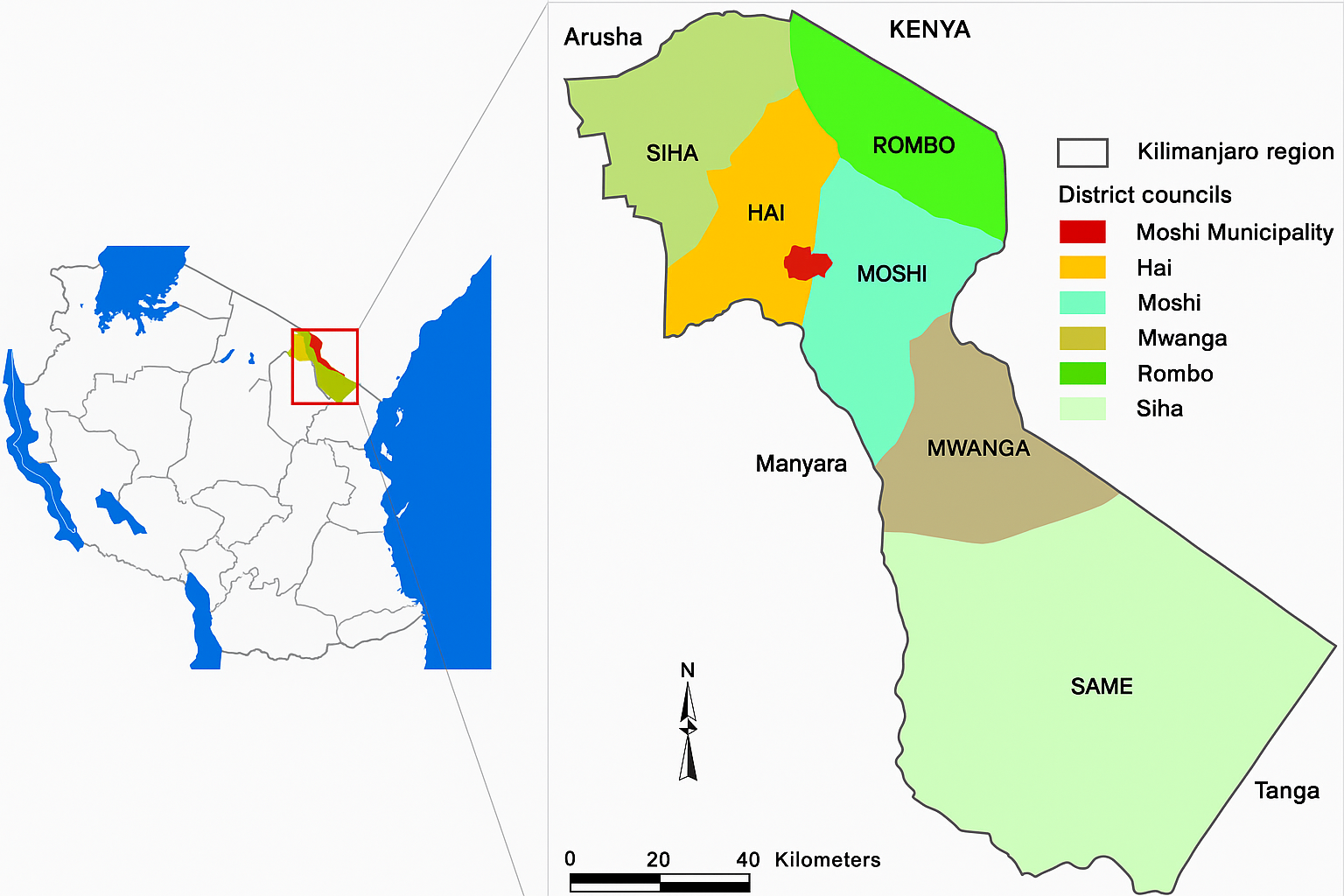

**Figure 2: Map of Moshi Municipality (Moshi Urban)** (18).

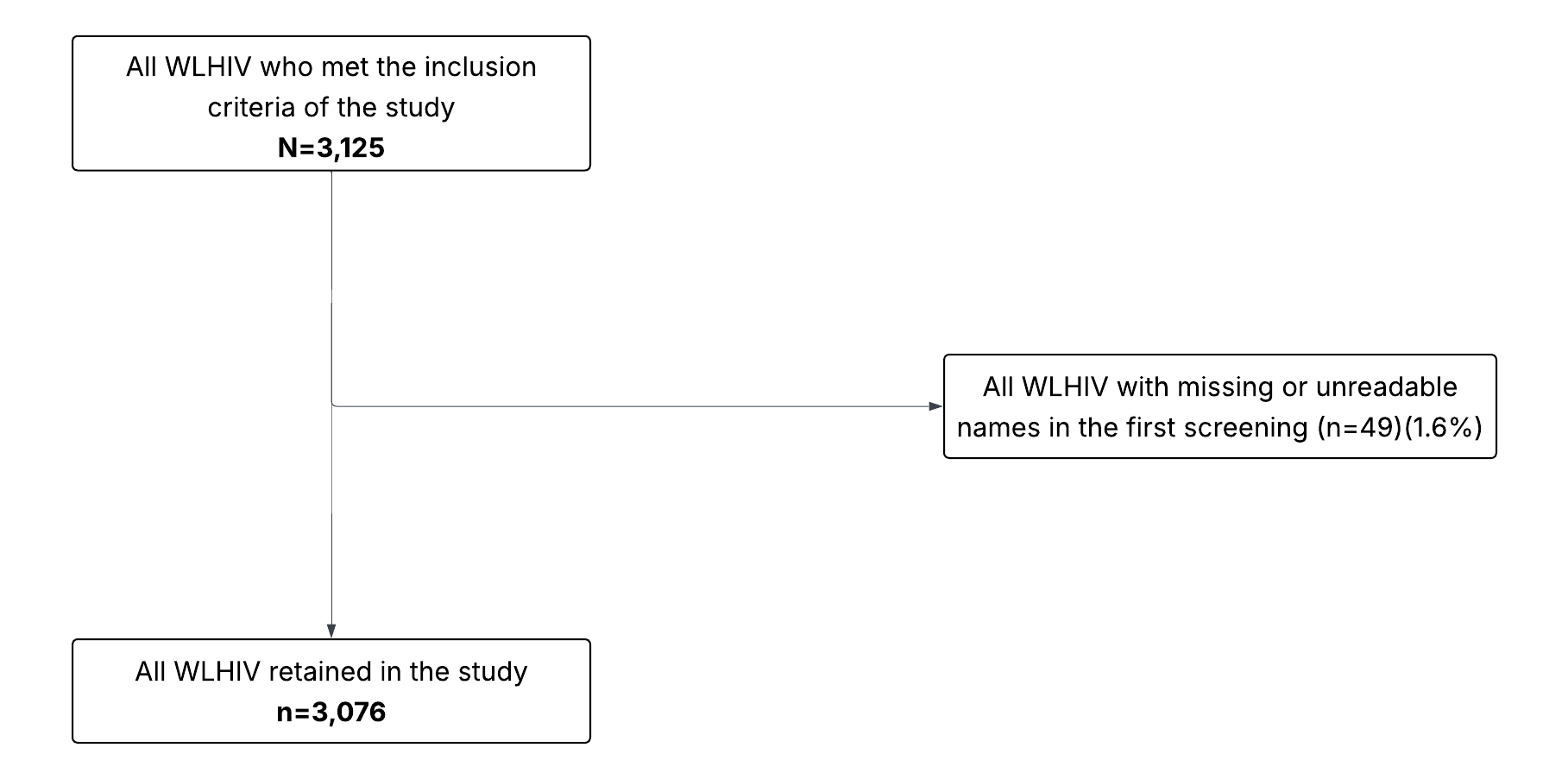

**Figure 3: Flowchart diagram for the selection of the study participants**
